# Leg-heel chest compression as an alternative for medical professionals in times of COVID-19

**DOI:** 10.1101/2021.03.09.21253220

**Authors:** Matthias Ott, Alexander Krohn, Laurence H. Bilfield, F. Dengler, C. Jaki, F. Echterdiek, T Schilling, J. Heymer

## Abstract

**Objective:** To evaluate leg-heel chest compression without previous training as an alternative for medical professionals and its effects on distance to potential aerosol spread during chest compression.

**Methods:** 20 medical professionals performed standard manual chest compression followed by leg-heel chest compression after a brief instruction on a manikin. We compared percentage of correct chest compression position, percentage of full chest recoil, percentage of correct compression depth, average compression depth, percentage of correct compression rate and average compression rate between both methods. In a second approach, potential aerosol spread during chest compression was visualized.

**Results:** There was no significant difference between manual and leg-heel compression. The distance to potential aerosol spread could have been increased by leg-heel method.

**Conclusion:** Under special circumstances like COVID-19-pandemic, leg-heel chest compression may be an effective alternative without previous training compared to manual chest compression while markedly increasing the distance to the patient.

## Introduction

SARS-CoV-2 is a highly contagious virus and causes COVID-19[1]. Its particles are mainly transmitted via droplets and aerosols[2]. The International Liaison Committee on Resuscitation (ILCOR) state chest compression having the potential to generate aerosol[3]. Likewise, the World Health Organisation (WHO) lists cardiopulmonary resuscitation (CPR) as an aerosol generating procedure (AGP)[4]. Distance is an important risk factor for virus transmission since the concentration of SARS-CoV-2 has been shown to diminish with increasing distance from a patient[2, 5, 6]. In some circumstances like bystander CPR, increased distance is the only adequate form of readily available protection against virus transmission. Therefore, we considered an alternative method for chest compression by leg-heel method for resuscitation. First described 1978 by Bilfield and Regula[7], the method has undergone further research for lay rescuers, professionals and school children[8–13]. As mentioned in ILCOR guidelines for resuscitation[3, 14], manikin studies showed similar results for leg-foot compression compared with standard manual chest compression[7, 9, 13]. We investigated whether medical professionals are able to provide adequate leg-heel chest compression on manikins with no more than very brief oral instructions. Secondly, we visualized potential aerosol spread towards the rescuer during leg-heel versus conventional manual chest compression.

## Methods

Medical professionals from either the emergency department or intensive care unit of Klinikum Stuttgart, Germany performed chest compression on a manikin (Resusci Anne Simulator, Laerdal Medical GmbH, Puchheim, Germany). Initially, they were told to perform standard continuous chest compression for 2 minutes to the best of their knowledge without any further instruction. After at least 5 minutes of recovery, the study coordinator gave each participant a one-minute lasting instruction on leg-heel chest compression by showing figures and reading out the original section from Bilfield and Regulas publication: Participants were told to remove the shoes, find the xiphoid process of the victim with the great toe and then moving the heel of the foot to a position 5 cm cranial to the xiphoid. They were told to stand directly over the victim with the length of the foot parallel to the sternum and with the weight-bearing foot planted at the side of the victim[7]. Without any previous training phase, they immediately performed 2 minutes leg-heel chest compression on the same manikin. Participants did neither get visual nor oral feedback on compression quality. Data was collected by SimPad PLUS (LLEAP Version 7.1.0.94, Laerdal Medical GmbH, Puchheim, Germany) and processed on a computer using Microsoft® Excel (Version 16.46, Microsoft Corporation, Redmond, WA, USA). The software recorded percentage of correct chest compression position, percentage of full chest recoil, percentage of correct compression depth (50-60mm), average compression depth in mm, percentage of correct compression rate and average compression rate. We applied Wilcoxon test to interpret data using Prism 9 for macOS (Version 9.0.2, GraphPad Software, San Diego, CA, USA).

To visualize potential aerosol spread during manual and leg-heel chest compression we used ultraviolet sensitive detergent as described previously[15]. Photos were taken by Nikon Df digital camera with a Sigma DG HSM Nikon Art 24.0 mm f/1.4 lens.

## Results

We recruited 20 trained nurses or physicians at the age of 29,3 ± 6,5 years. Mean body height was 1,74 ± 0,1m, mean body weight averaged 70,75 ± 15,2 kg. Calculated body mass index showed 23,17 ± 3,8 kg x m-2. Every participant completed both 2-minute cycles with no interruptions.

Table 1 shows data of manual compression compared to leg-heel compression. Percentage of correct chest compression position, of correct compression depth and the average compression depth was higher during manual compression. In contrast, percentage of full chest recoil and of correct compression rate was higher during leg-heel compression.

**Table 1:** Comparison of manual versus leg-heel comperisson during a two-minute period with n=20 participants ‡ correct comperssion depth was defined as 50-60mm.

|  | manual compression | leg-heel compression |
| --- | --- | --- |
| percentage of correct chest compression position | 95,3 ± 20,7 % | 80,0 ± 36,5 % |
| percentage of full chest recoil | 60,4 ± 39,3 % | 68,8 ± 33,3 % |
| percentage of correct compression depth‡ | 49,2 ± 40,6 % | 44,3 ± 38,4 % |
| average compression depth | 49,6 ± 8,6 mm | 46,8 ± 9,1 mm |
| percentage of correct compression rate | 36,8 ± 40,1 % | 42,6 ± 41,0% |
| average compression rate | 121,9 ± 15,9 min<sup>-1</sup> | 112,8 ± 19,2 min<sup>-1</sup> |

Results are also visualized in Figure 1. There is no significant difference between manual und leg-heel chest compression using Wilcoxon test on our results.

**Figure 1:**
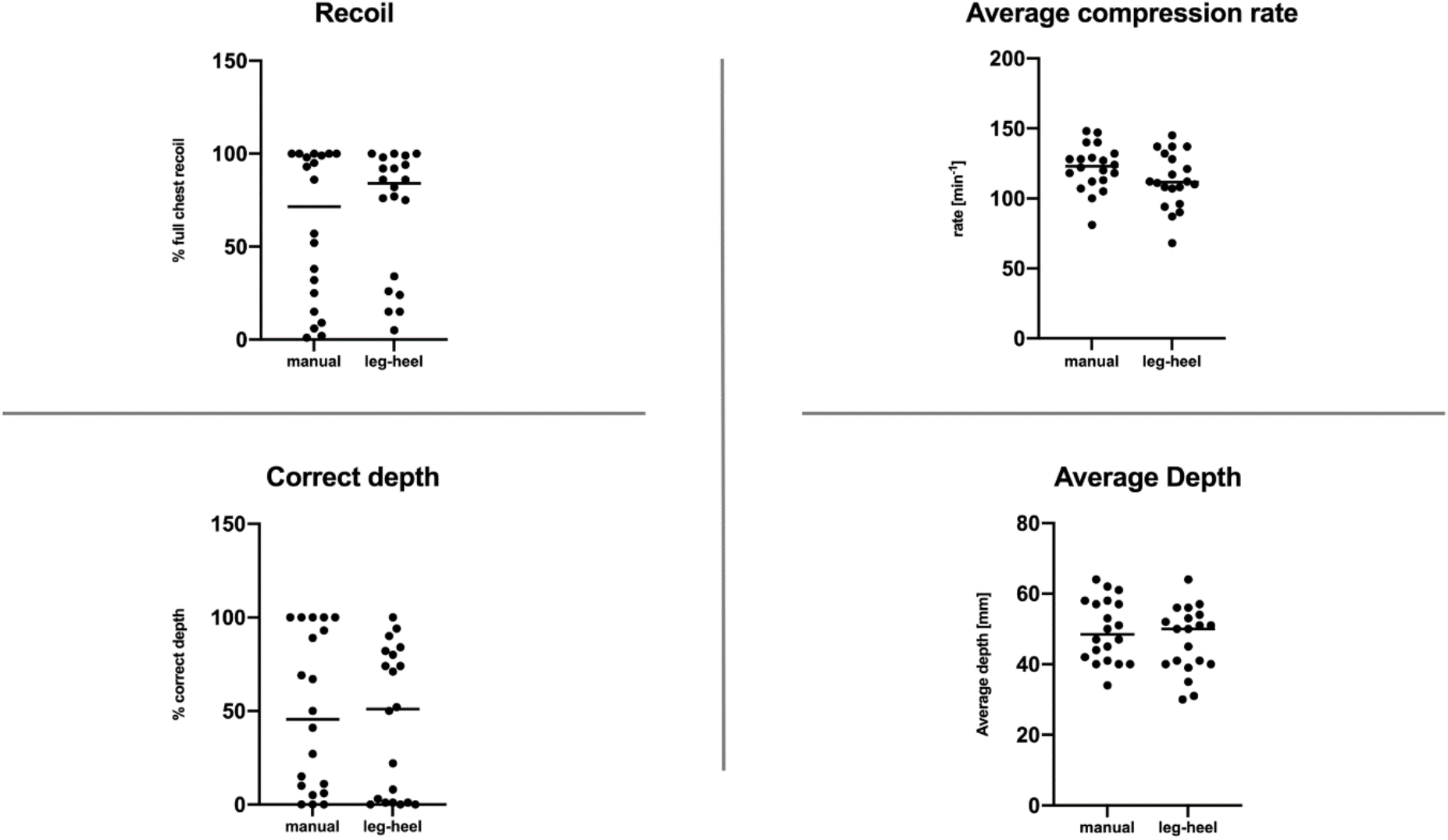
Results of Wilcoxon test on full chest recoil, average compression rate, average depth and percentage of correct compression depth.

Figure 2 and 3 showcase the 2-minute course of manual and leg-heel compression for two study participants. Chest compression rate and depth is illustrated in either yellow or green over time. Yellow areas signify compression rates <100 / >120 x min^-1^, compression depths <50 / >60mm or incomplete chest wall recoil. Green areas vice versa indicate adequate values.

**Figure 2:**
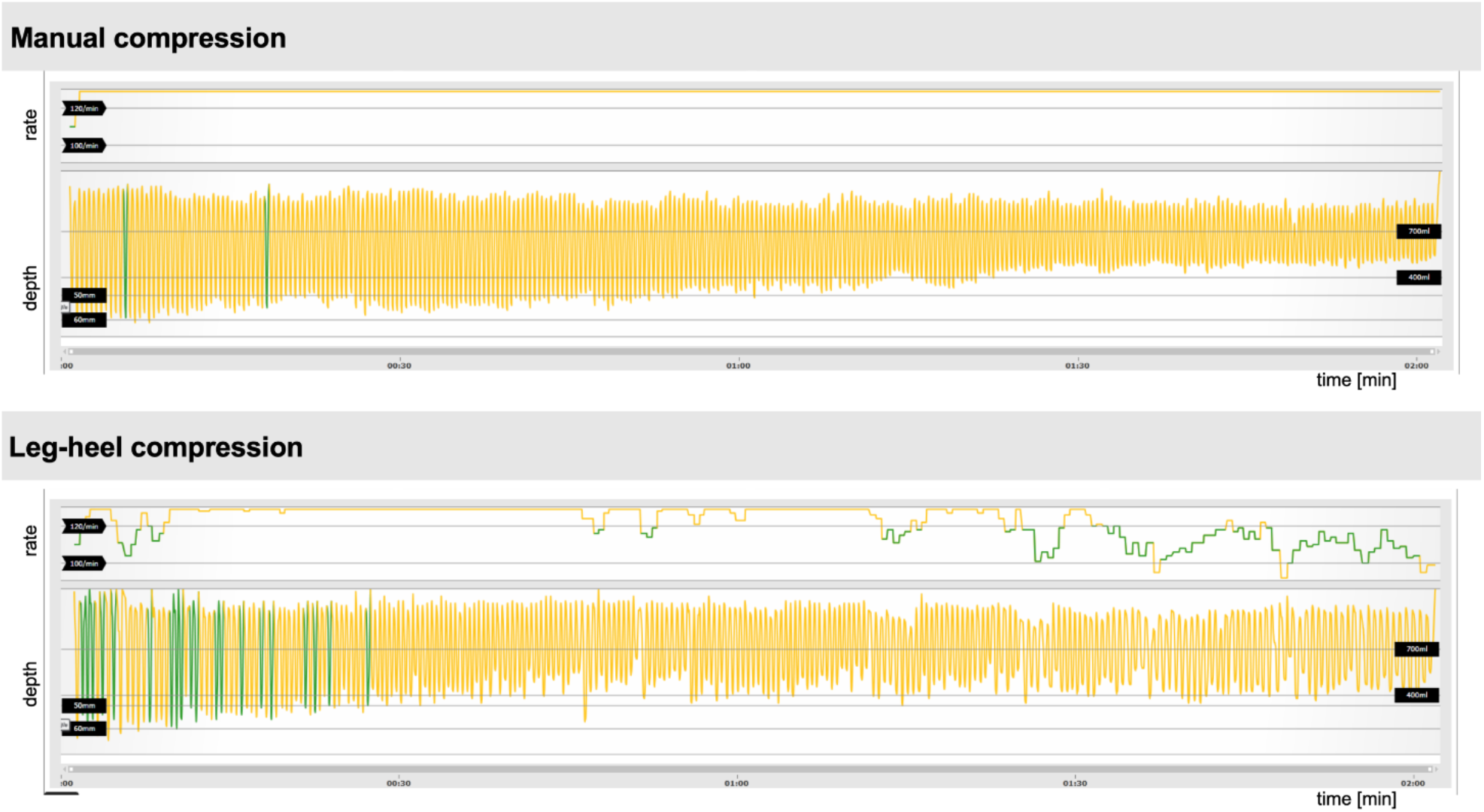
Participant A. Female, 1,67m body height, 52kg body weight, BMI 18,6kg x m^-2^

**Figure 3:**
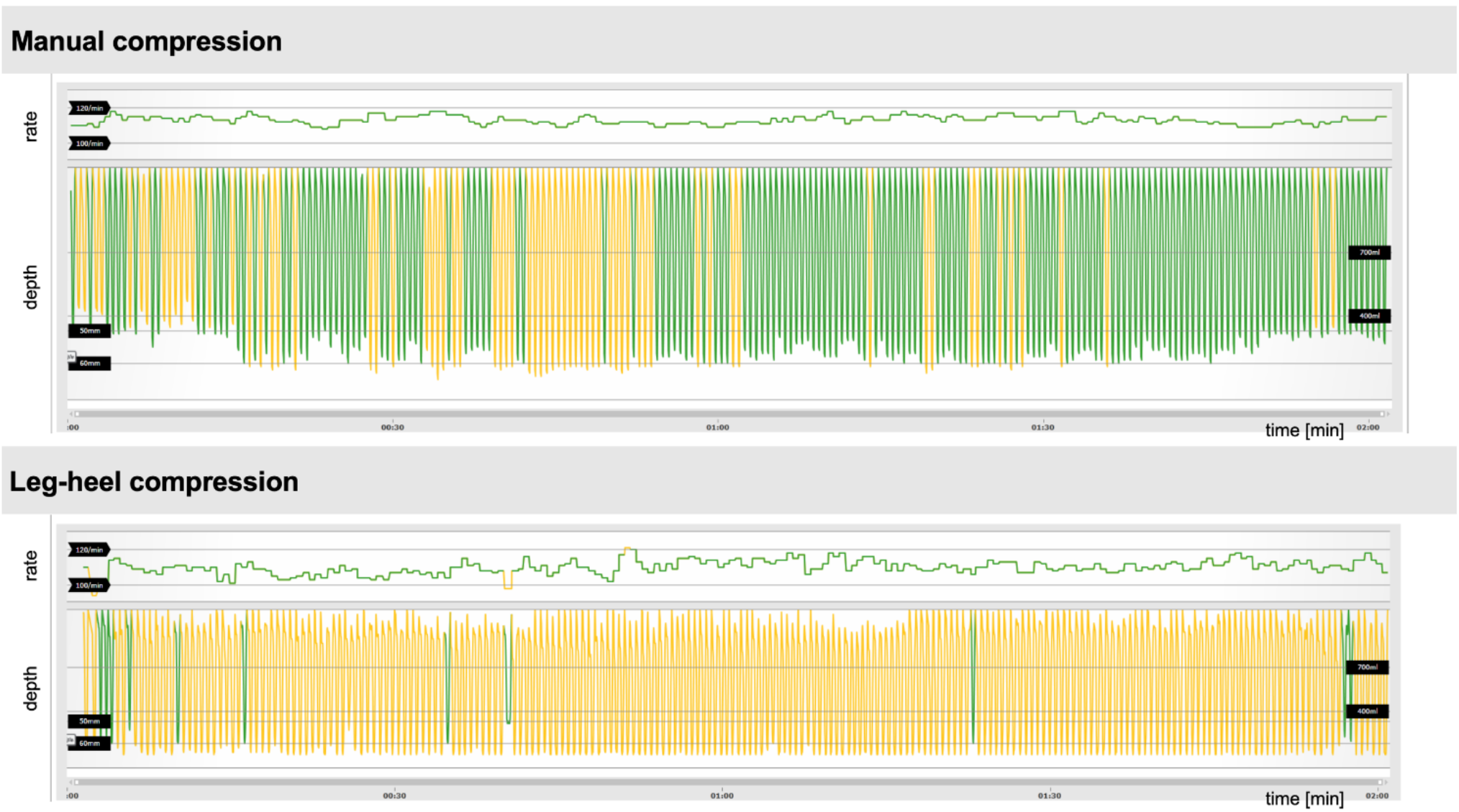
Participant B. Male, 1,88m body height, 81kg body weight, BMI 22,9 kg x m^-2^

Participant A applies more constant chest compression with more areas of adequate compression rate using leg-heel method.

Participant B generated more green areas using manual compression by full chest recoil and more adequate compression depth.

Figure 4 illustrates the results of the second part of our study. Visualized potential aerosol spread during manual chest compression on panel A is closer to the participants face than during leg-heel compression as illustrated on panel B.

**Figure 4:**
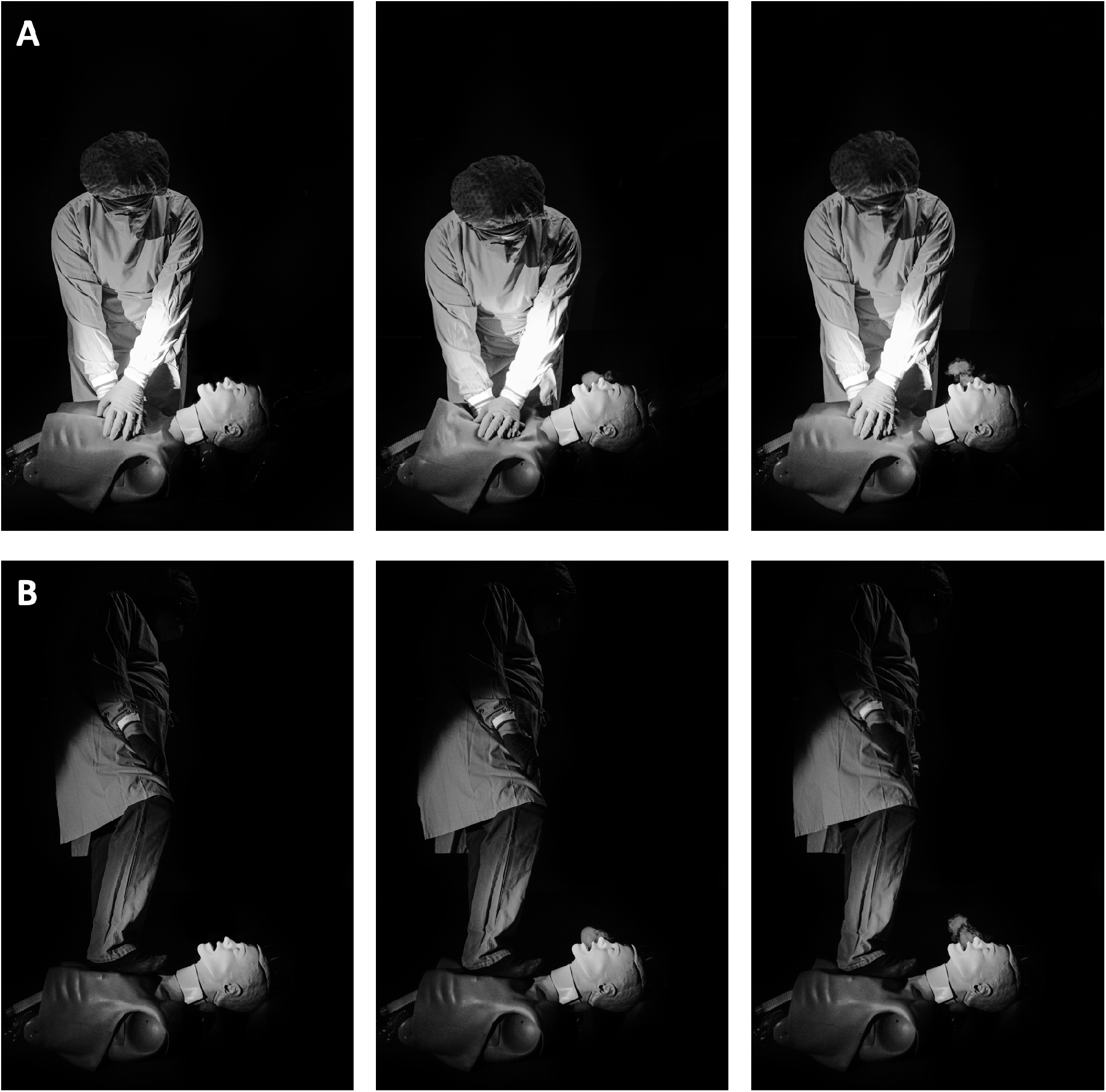
Visualized potential aerosol spread during manual chest compression (panel A) and leg-heel compression (panel B).

## Discussion

There is still no universal consensus which patient-related procedures carry an additional risk to generate aerosols[2]. Even a recently published review could not answer this question but states that aerosol generation is plausible during CPR[16]. However, many sources list cardiopulmonary resuscitation including chest compression as an AGP[1, 2, 4, 16–18]. Latest research on a swine cardiac arrest model also found significant aerosol generation during chest compression following defibrillation[19]. Potential effect on aerosol generation by ventilation during resuscitation stays unclear. However, in the face of a global pandemic with a highly contagious virus, ongoing shortages in personal protection equipment (PPE) and, in Europe, an increasing number of out of hospital cardiac arrest during the pandemic, prevention of infection risk for health-care professionals during CPR is of paramount importance[1].

Our data indicate no significant difference between manual chest compression and leg-heel chest compression regarding average compression depth, average compression rate, percentage of full chest recoil and percentage of correct compression depth. Because of the exploratory nature of this study and the limited number of participants we interpret our results as signals and consciously refrain from further statistical testing like non-inferiority testing was deliberately not performed. Previous manikin studies also showed no difference in depth and rate of chest compression when using the foot[7, 9, 13]. In contrast to our findings there are previous data indicating incomplete chest wall recoil during chest compression using the foot[7– 9]. This may lead to worse quality of CPR because complete chest wall recoil is important for hemodynamics during resuscitation and impacts patient outcome[20–23]. However, our findings trend to more complete chest recoil using leg-heel compression but comparing these findings with abovementioned studies is problematic because of differences in compression frequency, foot position and compression technique. Besides chest wall recoil, compression depth is also crucial for high quality chest compression[3, 10, 11, 21, 24]. Previous studies describe better results for compression depth when using foot compression especially for smaller, lighter or fatigued rescuers[7, 9, 10]. Our study cohort was too small for such subgroup analysis. Kherbeche et al. did not find advantage of foot compared to hand compression regarding compression depth for children and adolescents irrespective of body weight[8].

In contrast to published studies we only performed a 1-minute introduction to the method without intensive training. Interestingly, our results imply that this approach was not considerably worse than frequently trained manual chest compression and suggest that leg-heel compression may be a practical alternative for professionals under special circumstances. Besides clinical settings, leg-heel compression may also be an alternative for lay rescuers and bystanders not having PPE to their hands. Using leg-heel compression, they would be able to use their phone to call for help and furthermore. Likewise, Trenkamp and Perez described an increased ability to provide effective and uninterrupted compressions until arrival of help[10]. Considering the comparable outcomes after just a 1-minute introduction to the leg-heel compression method, further studies comparing outcomes after longer training of leg-heel compression are warranted. Leg-heel compression may also be an adequate alternative for fatigued rescuers.

The second part of our study revealed distinctly more distance to potential aerosol spread during chest compressions using the leg-heel compression method. Distance of rescuers to patient’s airway as a source of aerosols is a potential modifiable risk factor. In consequence of that, research of methods to increase this distance is urgently needed. This may also affect the willingness of bystanders to perform chest compression since preexisting fear of performing CPR may rise in times of COVID-19[25]. Our findings taken together suggest that leg-heel compression may be a practical alternative for CPR by emergency medical service during COVID-19 pandemic. Although infection risk during CPR still has unresolved issues, we think any form of prevention is still crucial until the COVID19-pandemic is over.

Limitations of our study are the limited number of participants and the limited comparability to previous studies due to different methods of foot compression. Hence, we feel the results of our study are primarily hypothesis generating and warranting further research on larger collectives. Another limitation may be that for safety precautions it is not feasible to put off shoes to apply chest compression, especially in out of hospital settings. However, we observed comparable results applying leg-heel compression with shoes on. Furthermore, overall chest compression quality in both methods was more or less surprisingly bad. Unlike other studies, we did not provide any kind of feedback to the participants which may partly explain our results. Manual chest compressions are basic skills for any medical professional and essential for patient outcome[3, 18, 21]. Once again, these findings underline the all-important role of regularly CPR trainings to lay rescuers as well as to medical professionals.

## Conclusion

Increased distance to patient’s airway is a readily available and well recognized form of protection from virus transmission, both for lay rescuers as well as medical professionals. Our results indicate that leg-heel chest compression may provide similar CPR quality compared to manual chest compression while markedly increasing the distance to the patients (potentially contagious) airway. In the light of the ongoing COVID-19-pandemic further evaluation of this alternative method of CPR appears promising.

## Data Availability

The datasets generated during and/or analysed during the current study are available from the corresponding author on reasonable request.

## Conflict of interests

None.

## Notes

### Competing Interest Statement

The authors have declared no competing interest.

### Funding Statement

No external fundings have been received.

### Author Declarations

There was no necessary IRB / ethics committee approval

